# Pro-arrhythmogenicity of single-tip vs. multi-spline catheters in post-MI VT ablation: a prospective, international, two-center experience

**DOI:** 10.64898/2026.08.06.26359917

**Authors:** Robert Rademaker, Maarten A.J. De Smet, Thomas Nia Jensen, Marta de Riva, Peter Lukac, Katja Zeppenfeld

## Abstract

**Background:** Substrate mapping using multielectrode catheters is increasingly used for post-myocardial infarction (MI) ventricular tachycardia (VT) avoiding repeated VT induction and mapping during VT. However, these catheters may mechanically induce ventricular arrhythmias with hemodynamic compromise. This study compares pro-arrhythmogenicity between single-tip and multi-spline catheters during functional substrate mapping.

**Methods:** Thirty post-MI patients (age 68±8 years, 97% male, LVEF 40% [IQR 33–46]) referred for VT ablation at two centers (2021–2024) underwent endocardial mapping during baseline rhythm in random order with both a multi-spline catheter (Octaray™, n=4; Pentaray™, n=26) *and* a single-tip QDOT™ catheter. The protocol was prematurely terminated if (i) two mechanically induced VTs required ECV, (ii) recurrent ATP-treated mechanical VTs caused hemodynamic compromise, or (iii) excessive mechanically induced ectopy impaired catheter contact. Mapping time, point density, and mechanically induced arrhythmias were assessed.

**Results:** Multi-spline catheters enabled faster mapping (26±9 vs 60±16 minutes, p<0.001) with more acquired points (p<0.001). VTs were more frequently mechanically induced with multi-spline catheters (median 2 [IQR 1–4] vs 0 [0–3], p<0.05) and these VTs were faster (304ms, IQR 292–320] vs 373ms, IQR [316–405], p=0.01) and degenerated more often into VF (3 vs. 0). Overall, 17 patients (57%) experienced at least one mechanically induced VT; seven (23%) required cardioversion, and mapping was prematurely terminated in eight (27%), all while using multi-spline catheters.

**Conclusion:** Multi-spline catheters allow rapid substrate mapping but with substantial risk of mechanically induced arrhythmias, requiring premature termination of substrate mapping because of safety concerns. Their use in post-MI VT ablation warrants careful risk–benefit assessment.

**Clinical perspectives:** *What is known:*

◦ Substrate mapping using multielectrode catheters is increasingly used for post-myocardial infarction (MI) ventricular tachycardia (VT) avoiding repeated VT induction and mapping during VT.
◦ However, these catheters may mechanically induce ventricular arrhythmias with hemodynamic compromise.

*What this study adds:*

◦ This is the first prospective, systematic comparison of incidence and characteristics of catheter induced ventricular arrhythmias between multi-spline and single-tip catheters during functional substrate mapping for post-myocardial infarction VT.
◦ Multi-spline catheters were associated with a higher incidence of mechanically induced ventricular arrhythmias, including ventricular fibrillation, requiring premature termination of substrate mapping in more than a quarter of patients because of safety concerns.
◦ Mapping with multi-spline catheters in VT-ablation warrant a careful risk-benefit assessment due to possible safety concerns over mechanically induced arrythmias.

**Graphical abstract:** 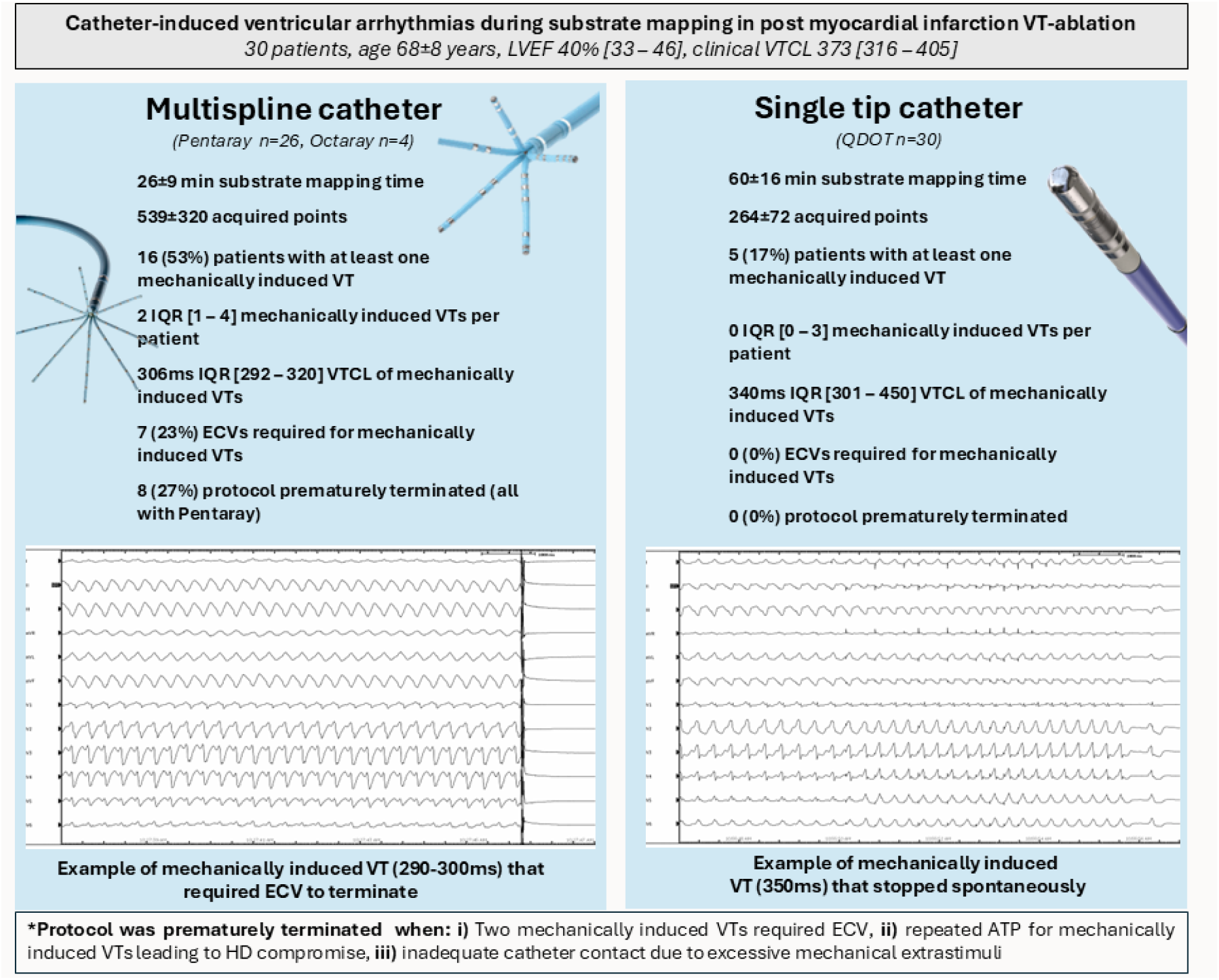

## Introduction

Substrate mapping in ventricular tachycardia (VT) ablation is increasingly performed. It may improve patient safety by avoiding repeated VT induction and mapping during VT potentially causing hemodynamic compromise. (1)The use of multielectrode, multispline catheters has been suggested for endocardial mapping of post-myocardial infarction (MI) VT as they may allow for higher mapping point density, better near-field resolution, and higher procedural efficiency. (2–8).

However, catheter-induced ventricular arrhythmias (VAs) pose significant challenges during substrate mapping. When sustained, their termination requires antitachycardia pacing (ATP) or electrical cardioversion (ECV), which carry the risk of map shifts, hemodynamic instability, and even decompensated heart failure. (9) Furthermore, frequent catheter-induced ectopy can compromise mapping accuracy and hinder systematic substrate mapping. Consequently, repeated mechanically induced VAs may negate the safety and efficiency benefits of multi-electrode mapping.

While informal discussions among electrophysiologists have raised concerns about the pro-arrhythmic effects of multispline catheters, no systematic evaluation of the incidence, characteristics and consequences of mechanically induced VAs has been conducted. This study aimed to assess and compare the mapping efficiency and mechanical pro-arrhythmic effects of multispline and single-tip catheters during functional substrate mapping in patients undergoing ablation for post-MI VT.

## Methods

### Data availability statement

All data and analytical methods supporting the findings of this study are available from the corresponding authors upon reasonable request.

### Patient population

Consecutive patients with a history of MI undergoing VT ablation at Leiden University Medical Center (LUMC) and Aarhus University Hospital (AUH) between June 2021 and March 2024 were eligible for the study. Those who provided informed consent were included. Ethical approval was granted by the institutional review boards (NL 74386.058.20), and written informed consent was obtained from all participants. The study adhered to Good Clinical Practice standards, institutional protocols and the Declaration of Helsinki.

### Pre-ablation assessment

Prior to the procedure, all participants underwent a comprehensive evaluation. Clinical records were reviewed for ischemic events, reperfusion procedures, prior revascularization (PCI or CABG), management of heart failure, and anti-arrhythmic drugs. Ischemia was assessed with stress testing, coronary angiography, or CT; significant lesions were revascularized before ablation. Presenting VTs were reviewed for cycle length (CL) and method of termination based on ICD interrogation and/or 12 lead ECG review. Additionally, VTs were classified as hemodynamically stable or unstable. VTs were considered hemodynamically unstable if acute electrical cardioversion was required and/or if severe symptoms (e.g., syncope, presyncope, angina, or dyspnea) were reported. All other VTs were classified as stable. Echocardiography was conducted to rule out left ventricular (LV) thrombus and to measure LV function using Simpson’s biplane method. Patients underwent a pre-procedural contrast-enhanced, ECG-gated CT scan, with 3D segmentation of cardiac anatomy and wall thickness performed using ADAS 3D software (ADAS3D Medial SL, Barcelona, Spain).

### Mapping protocol

Mapping and ablation were carried out under conscious sedation, deep sedation, or general anesthesia, at the discretion of the operator. Antiarrhythmic therapy, except amiodarone, was withheld for a minimum of five half-lives. At the beginning of the procedure, programmed electrical stimulation (PES) was performed using up to four drive CL (600, 500, 400, and 350 ms), and up to four extrastimuli (down to 200 ms or the ventricular refractory period (VRP)) at two right ventricular (RV) and at least one LV site to determine the VRP and to induce VT. A VT was considered clinical if its 12-lead morphology and cycle length (Δ <30 ms) matched a previously documented VT, either on 12- lead ECG or based on ICD interrogation. Fast VT was defined as VTCL ≤ VRP+30ms. (10)

Electroanatomic mapping (EAM) of the LV was performed during sinus rhythm, RV pacing, and after a single short coupled right ventricular extrastimulus (RVE) using CARTO3™ (Biosense Webster). CT-derived anatomy was aligned with the EAM using the left main coronary artery, aorta, and LV apex as landmarks. Detailed substrate mapping was focused on areas with wall thinning on the CT scan using both the QDOT^TM^ and a multi-spline catheter (PENTARAY™ or OCTARAY^TM^ catheters), with the initial catheter chosen randomly. The mapping time and number of acquired points for each map was noted. Sites with evoked delayed potentials (EDP) were tagged on the maps. During the procedure, the stimulator and EP recording system (Prucka, GE Healthcare) were operated by an experienced EP fellow or electrophysiologist. In case of VT induction (mechanically or by electrical stimulation), long bursts of ATP starting with a CL 10% shorter than the VTCL were applied to terminate the VT. Only after failed ATP attempts or in case of severe hemodynamic compromise, ECV was used. For each induced VT, the VTCL and method of induction and termination were noted. Per protocol, the mapping procedure was prematurely terminated if any of the following occurred: (i) two episodes of mechanically induced VTs requiring ECV, (ii) repeated ATP for mechanically induced VTs leading to hemodynamic compromise, or (iii) excessive mechanically induced extrastimuli hampering adequate catheter contact. Ablation was performed after completion of substrate mapping using the Qdot catheter aiming for EDP elimination. Radiofrequency energy was delivered at 40–50 W (temperature cap 43°C, irrigation 20–30 mL/min, impedance drop >10 ohms) until high-output pacing (10 mA, 2 ms) failed to capture. (11, 12) Further mapping and ablation was performed with the Qdot catheter if sustained monomorphic VT (SMVT) remained inducible. Only fast VTs induced after substrate modification were not further targeted.

### Procedural outcomes

Acute ablation outcome was classified as: (1) no inducible SMVT, (2) inducible clinical VT (matching pre-ablation morphology) or (3) inducible non-clinical VT. Patients non-inducible both before and after ablation were categorized as non-inducible. Success was defined as absence of inducible SMVTs and elimination of EDPs confirmed by re-mapping.

### Post-ablation care and follow-up

Echocardiography was repeated post-procedure to assess LV function and exclude pericardial effusion. Patients without ICDs received implantation per guideline recommendations. (13) Antiarrhythmic therapy was maintained until first follow-up. Scheduled visits occurred at 3 months and then every 6 months. Recurrence was defined as any VT captured by the ICD or on 12-lead electrocardiogram lasting >30 s or requiring ICD therapy. The CL of clinical VTs before ablation, procedural VTs and recurrent VTs were compared.

### Statistical analysis

Continuous data were expressed as mean ± standard deviation or median with interquartile range [Q1–Q3] and compared using the Student’s t-test or Mann–Whitney U test as appropriate. Categorical variables were reported as counts and percentages, analyzed with chi-square or Fisher’s exact test. Statistical significance was set at a two-tailed P-value <0.05. Analyses were conducted in SPSS v27.0 (IBM).

## Results

### Patient characteristics

Thirty post-MI patients (mean age 68±8 years, 97% male, median LVEF 40% [IQR 33-46]) undergoing VT ablation at Leiden University Medical Center (n=27) and Aarhus University Hospital (n=3) were prospectively included. In 87% of the patients, it was the first VT ablation. Eighteen patients (60%) had an ICD implanted and 13% were on amiodarone. Sixteen patients (53%) had undergone prior PCI and 12 (40%) prior CABG.

The median VT burden in the six months prior to the procedure was 3 episodes IQR [1 – 4] per patient, with a median clinical VTCL of 373ms IQR [316 – 405]. The presenting VT was hemodynamically tolerated in eight patients (27%). Clinical VTs were terminated by ATP in 20%, ICD- shock in 40%, external ECV in 20%, anti-arrhythmic drugs in 13%, and stopped spontaneously in two patients (7%). See Table 1 for details.

**Table 1.**
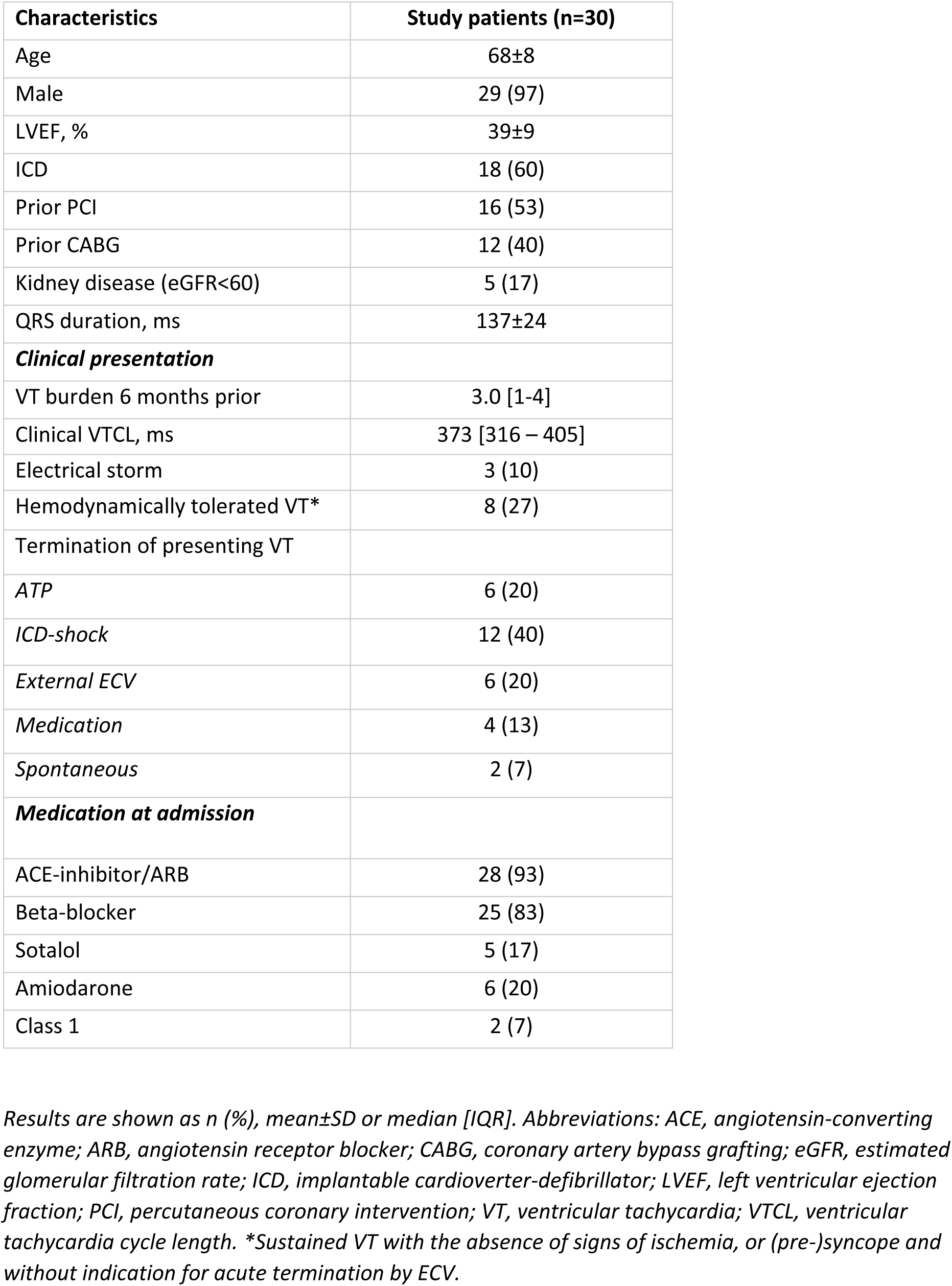
Baseline characteristics.

### Procedural data and mapping times

Mapping and ablation were performed in conscious sedation in 25 patients (83%), deep sedation in two patients (7%) and general anesthesia in three patients (10%). At the beginning of the procedure, 27 (90%) patients were inducible for any VT with a median VTCL of 330 ms IQR [298 – 420]. The (presumed) clinical VT was induced in 22/30 (73%) patients. The median VTCL of induced clinical VTs was 335ms IQR [306 – 426]. The median VTCL of induced, non-clinical, VTs was 250ms IQR [244 – 413].

Two separate functional substrate maps were attempted in a random order with a multi-spline catheter (Octaray^TM^ (n=4) or Pentaray^TM^ (n=26)) and the single-tip QDOT^TM^ catheter focusing on the area of wall thinning determined by preprocedural cardiac CT. The multi-spline catheters were used to create the first map in 11/30 cases (37%). Mean total procedural time was 299±53 minutes. Total substrate mapping time was 85±22 minutes with significantly longer mapping times for the single-tip catheter as compared to multi-spline catheters (60±16 minutes vs 26±9 minutes respectively, p-value <0.001). Substrate maps with multi-electrode mapping catheters had significantly more mapping points compared to single-tip, both for sinus rhythm (539±320 vs 264±72 mapping points, p- value<0.001) and functional substrate maps (148±51 vs 74±27, p-value<0.001) maps. Complete sampling of the endocardium was achieved with fill threshold <10mm when mapping with the QDOT catheter. See Table 2 for details.

**Table 2.** Procedural data.

|  | **Study patients (n=30)** |
| --- | --- |
| Procedural time, min | 299±53 |
| Substrate mapping time, min | 85±22 |
| Mapping protocol prematurely terminated* | 8 (27%) |
| ***VT inducibility*** |  |
| Inducible for VT before ablation | 27 (90%) |
| VTCL VT inducible before ablation, ms | 330 [298 – 420] |
| Patients with at least one mechanically induced VT | 17 (57%) |
| Number of mechanically induced VTs per patient | 2 [1 – 6] |
| VTCL mechanically induced VTs multispline catheter, ms | 306 [292 – 320] |
| VTCL mechanically induced VTs single tip catheter, ms | 340 [301 – 450] |
| Patients with ≥1 required ECV for mechanically induced VTs | 7 (23%) |
| Inducible for any VT after ablation | 8 (29%) |
| Clinical VT inducible after ablation | 1 (4%) |
| Only fast-VT inducible after ablation** | 5 (18%) |
| VTCL induced VTs after ablation, ms | 253 [237 – 383] |

Results are shown as n (%), mean±SD or median [IQR]. See Table 1 for abbreviations.
\*(i) two distinct mechanically induced VTs requiring ECV, (ii) repeated ATP for mechanically induced VTs leading to hemodynamic compromise, or (iii) inadequate catheter contact due to excessive mechanically induced premature ventricular contractions
\*\*VTCL= RV refractory period +<30ms

### Acute procedural outcome

In two patients, induction after ablation was not performed: in one due to persistent inducibility of the clinical VT with an unreachable substrate, and in the other because the patient could no longer tolerate the procedure performed under conscious sedation. After ablation, 20/28 patients (71%) were non-inducible from the RV and multiple LV sites. In eight patients (29%), any SMVT remained inducible (median VTCL 253ms IQR [237 – 383], but in only one patient the clinical VT remained inducible (VTCL 330ms). Of the remaining seven patients, five were only inducible for fast VTs (mean VTCL 242ms) and two for VTs not earlier observed spontaneously (VTCL 400/420ms). See Table 2 for details.

### Mechanically induced ventricular arrhythmias

In 17 patients (57%), a total of 66 sustained VA were mechanically induced during mapping (median 2 VA per patient, IQR 1-6). Of those, 63 were SMVT and three polymorphic VT degenerating into VF. All three PVT/VF episodes occurred in three different patients while mapping with the Pentaray^TM^ catheter. There was a significantly higher number of mechanically induced VTs with the multi-spline catheters compared to the single-tip (median induced VTs 2 IQR [1 – 4] versus median 0 IQR [0 – 3], p-value<0.05). Seven patients (23%) required at least one ECV due to mechanically induced VA (SMVT/ventricular fibrillation) and three patients (10%) required two ECVs.

In eight (27%) patients, the protocol was prematurely terminated because of recurrent ECVs (3/8), repeated ATP with hemodynamic compromise (1/8) and unacceptable number of catheter-induced ectopy precluding adequate catheter contact (4/8). All required ECV and premature termination of the protocol occured while mapping with the Pentaray^TM^ catheter. See Table 2 and the Graphical Abstract for details.

### VT characteristics comparison

The VTCL of the clinical VT, the VT induced by PES before ablation and the VTs mechanically induced by the single tip catheter were similar (373ms IQR [316 – 405], 330ms IQR [298- 420, and 340ms IQR [301 – 450], respectively, one-way ANOVA p=0.88)). In contrast, the VTs mechanically induced by the multi-spline catheters had a significantly shorter cycle length compared to the clinical VT (304ms IQR 292 – 320] versus 373ms IQR [316 – 405], p=0.01).

After ablation and completing the entire PES protocol (down to a drive train of 350ms with up to 4 extrastimuli), the VTCL of the remaining inducible VT was shorter (median VTCL 253ms [237–383]). See Figure 1 for details.

**Figure 1.**
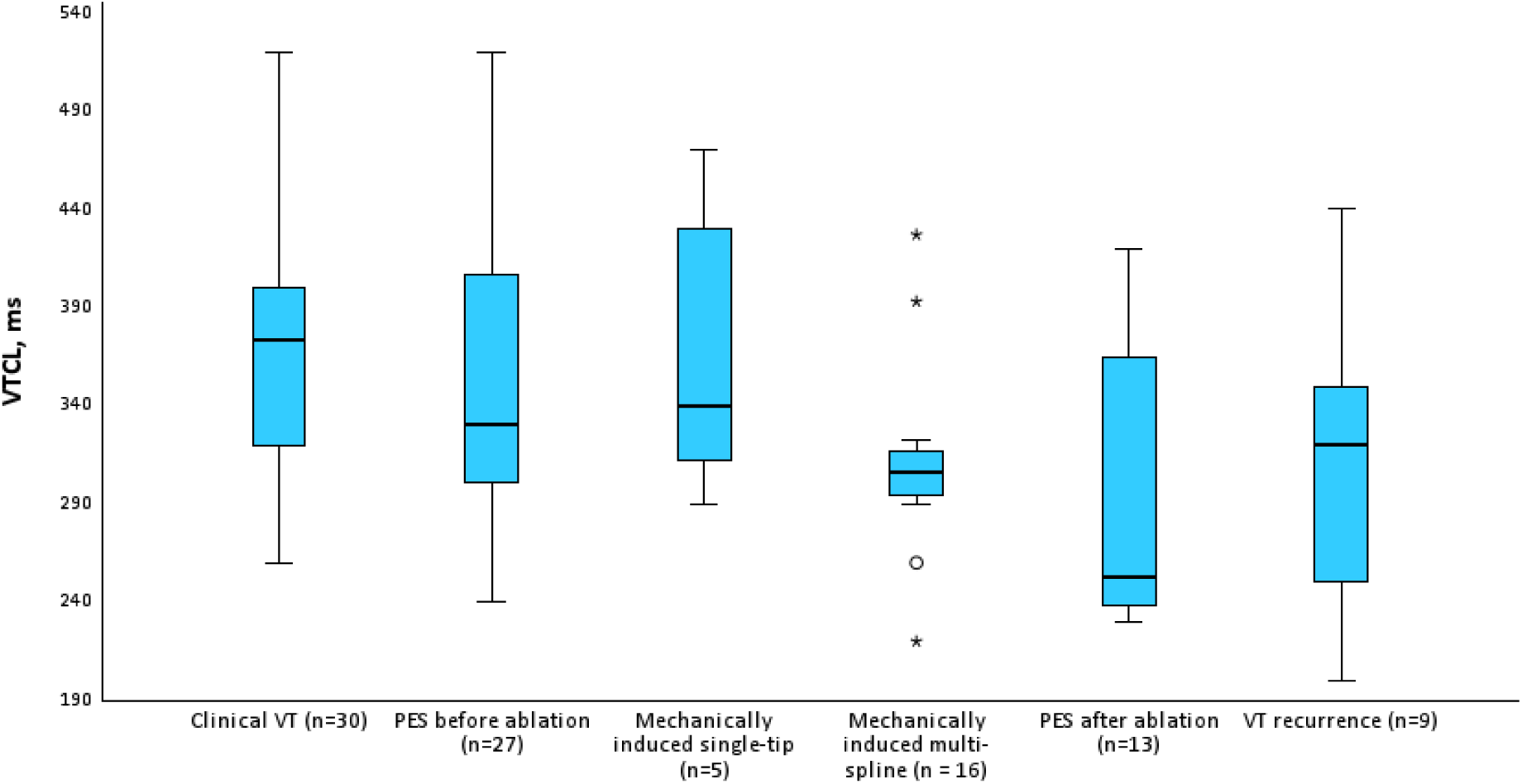
Boxplot showing induced VTCL stratified by induction. PES, programmed electrical stimulation; VT, ventricular tachycardia

### Follow-up

The mean follow-up time after ablation was 21±13 months. During follow-up, 9 patients (30%) experienced VT-recurrence (median recurrence VTCL 320ms IQR [235 – 354] after a median of 6 months IQR [5 – 24]. VTs occurring during follow-up were numerically faster than the presenting clinical tachycardias (320ms IQR [235 – 354] versus 373ms IQR [316 – 405], p=0.14).

All patients with recurrence had an ICD implanted at that time. Four out of nine patients required an ICD shock to terminate the VT, four required ATP, and in one patient the tachycardia terminated spontaneously. Three patients died during follow-up (lung cancer, terminal heart failure, and respiratory insufficiency caused by pneumonia). See Table 3 for details.

**Table 3.** Follow-up data.

|  | Study patients (n=30) |
| --- | --- |
| Follow-up, months | 21±13 |
| VT recurrence | 9 (30) |
| VTCL recurrence | 320 [235 – 354] |
| ATP termination | 4/9 |
| ICD shock | 4/9 |
| Spontaneous termination | 1/9 |
| Death during follow-up | 3 (10) |

Results are shown as n (%), mean±SD or median [IQR]. See Table 1 for abbreviations.
ATP, anti-tachycardia pacing; ICD, internal cardiac defibrillator

## Discussion

### Main Findings

This prospective, two-centre study of patients undergoing post-MI VT functional substrate mapping is the first to systematically compare the incidence and characteristics of mechanically induced VAs using multi-spline mapping catheters versus a single-tip contact force-sensing catheter.

Mapping of the target area was significantly faster and achieved a higher point density using multi-spline catheters. However, this approach was associated with a higher incidence of mechanically induced VA which had a shorter cycle length.

Almost one-quarter of patients required ECV due to catheter-induced VA during the procedure, and in 27% of cases, the mapping protocol had to be prematurely terminated due to recurrent mechanical VA. Notably, these events occurred exclusively during multi-spline catheter mapping.

Together, these findings highlight a trade-off between mapping efficiency and safety. While multi-spline catheters offer superior mapping efficiency, they also increase the risk of mechanically induced arrhythmias, which can compromise procedural safety.

### Substrate mapping and procedural safety

Changing morphologies, hemodynamic instability, and non-inducibility of VT excludes VT activation mapping as a reliable method for identifying ablation target sites. (14, 15)

Even when stable monomorphic VT can be induced, repetitive VT induction, prolonged mapping during initially stable VT, and repeated cardioversions may significantly compromise patient safety.

While general anesthesia improves patient comfort during the procedure, in particular if VTs are frequently induced and/or repeated cardioversions are required, it also prolongs the procedure and reduces hemodynamic tolerance to induced VTs. In the present study, the majority of patients underwent the procedure under conscious sedation, which may favorably influence hemodynamic tolerance during mechanically induced VTs by preserving physiological compensatory mechanisms compared with general anesthesia or deep sedation.

Substrate mapping without the need for (repeated) VT induction has been proposed to not only increase procedural success, but also procedural safety. (11, 16, 17). Multi-spline, multi-electrode catheters have been used for endocardial ventricular substrate mapping allowing for a higher mapping point density, higher procedural efficiency and, dependent on electrode size and spacing, improved near-field resolution. The efficiency of multi-electrode catheters has been well documented in atrial mapping, where higher density maps and reduced acquisition times translate into more efficient workflows (18, 19). Our study confirms that detailed substrate mapping, including pacing maneuvers for functional substrate mapping, can be performed in less than 30 mins with high mapping point density of the area of interest using multi-spline catheters, provided that the map can safely be completed. However, in contrast to the atria, the complex 3D anatomy of the endocardial left chamber with its intracavitary structures (e.g. papillary muscles, chordae) requires more complex catheter manipulation, which may explain the high incidence of mechanically induced VA .

Of concern, these mechanically induced VA were faster, included polymorphic VT and VF, and required more often ECV for termination. Although mechanically induced SMVT also occurred occasionally during mapping with a linear, single-tip catheter, these VTs were slower and could be terminated by ATP. The pro-arrhythmic effects of multi-spline catheters are likely multifactorial. The flexible splines make contact with multiple adjacent endocardial sites, increasing the risk of mechanically depolarizing the myocardium. This may lead to repetitive premature ventricular complexes, the induction of scar-related macro re-entrant VTs, as well as repolarization dispersion, which may trigger polymorphic VT and localized re-entry with shorter VTCL.

Of note, in 14% of our cohort the mapping with multi-spline catheters has been prematurely terminated not because of safety issues but because of an inacceptable high burden of catheter induced ventricular ectopy which precluded systematic substrate mapping and the application of the functional substrate mapping protocol.

### Limitations

Several limitations of our study must be acknowledged. The study included only 30 patients, however we terminated the study due to safety concerns due to frequent required cardioversion. The findings may not be generalizable beyond the post-MI population, particularly to non-ischemic cardiomyopathy, where substrate characteristics differ. Although grouped as “multi-spline,” Octaray™ and Pentaray™ differ in design. The fact that polymorphic arrhythmias were only observed with the Pentaray™ may suggest device-specific effects that cannot be fully disentangled in this sample. The potential impact of multi-electrode mapping on ablation outcome cannot be evaluated, since 73% of patients underwent the full mapping protocol with both catheter-types.

### Future directions

Larger, multicenter studies are warranted to confirm the proarrhythmic effects of multi-spline catheters across patient subgroups. Advances in catheter engineering, including material and design of novel multi-electrode catheters may reduce the risk of mechanically induced VA. However, development of new catheters for substrate mapping in VT should also encompass a rigorous assessment of mechanical pro-arrhythmic risk for the different cavities.

## Conclusions

Multi-spline catheters allow rapid substrate mapping but with substantial risk of mechanically induced arrhythmias, requiring premature termination due to safety concerns. Their use in post-MI VT ablation warrants careful risk–benefit assessment.

## Data Availability

Data will be made available upon reasonable request to the corresponding author

